# Global solidarity in genomic surveillance improves early detection of respiratory virus threats

**DOI:** 10.1101/2023.11.01.23297901

**Authors:** Simon P.J. de Jong, Brooke E. Nichols, Anniek de Ruijter, Edyth Parker, Vera Mitesser, Christian Happi, Menno D. de Jong, Alvin X. Han, Colin A. Russell

## Abstract

Public health decision-making for respiratory virus outbreaks relies heavily on genomic sequencing to detect new (variant) viruses. However, respiratory virus sequencing infrastructure is highly unequally distributed globally, potentially limiting the efficiency and effectiveness of surveillance efforts and raising concerns about preparedness for future threats. Using mathematical models, we demonstrate that relative to global sequencing efforts during the COVID-19 pandemic, increased global solidarity in respiratory virus genomic surveillance would vastly improve the capacity to rapidly detect novel threats, even with a substantially reduced number of viruses sequenced globally, leading to improved effectiveness and efficiency. As a result, the time between a (variant) virus’ first global detection and first local case would increase in all countries, allowing for more time to design and implement global and local public health measures to mitigate the threat’s potential public health impacts. Our results show that operationalizing global health solidarity is key to guiding investment in preparedness for future pandemic threats.

## Introduction

Genomic surveillance of respiratory viruses is a critical component of public health preparedness and response, particularly for identifying and monitoring the spread of new viruses and their variants^1–3^. According to Article 5 of the International Health Regulations (IHR), Member States to the World Health Organisation (WHO) are obligated to ensure national surveillance capacity^4^. The COVID-19 pandemic represented the zenith of global respiratory virus sequencing output so far, with ∼7 million SARS-CoV-2 genomes submitted to GISAID (www.gisaid.org) in 2022 alone. However, this output was highly unequally distributed^5^: half of all publicly shared genomes originated from countries that account for only 4.4% of the global human population while half of the global population accounted for only 0.7% of publicly shared genomes (Fig. 1a, Extended Data Fig. 1). Novel viruses and their variants can potentially emerge in any country. As a result, the unequal distribution of sequencing infrastructure potentially strongly limits the global capacity to rapidly detect novel threats.

**Figure 1.**
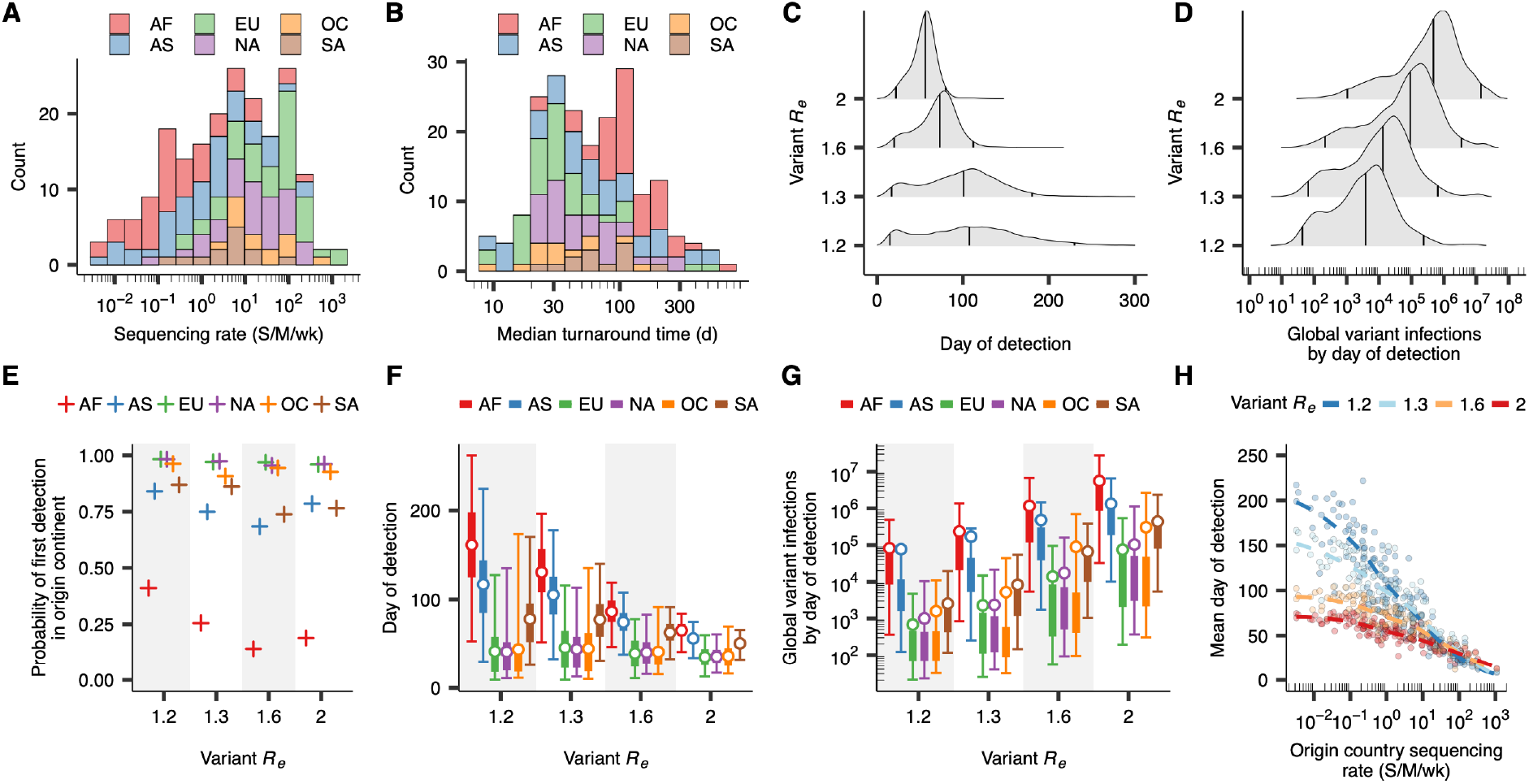
The global time to variant detection based on the SARS-CoV-2 genomic sequencing landscape in 2022. **(A)** Distribution of non-zero country-specific weekly sequencing rates per million people by continent estimated from GISAID metadata (*N* = 199) (AF: Africa, EU: Europe, OC: Oceania, AS: Asia, NA: North America, SA: South America). **(B)** Distribution of median country-specific time from sample collection to sequence deposition in GISAID, i.e. turnaround time (*N* = 199). **(C)** The distribution of days to variant detection for different values of variant *R*_*e*_ in global metapopulation model simulations, each with a distinct scenario of variant emergence *(N* = 10,000 for each variant *R*_*e*_). Vertical lines correspond to the median and 95% CI. **(D)** The simulated distribution of the number of global variant infections by the day of first variant detection. **(E)** The simulated probability that the variant is first detected in its origin continent, by origin continent. **(F)** The simulated time to variant detection by variant origin continent. Thin and thick lines correspond to 95% and 50% CIs, respectively. Points correspond to means. **(G)** The simulated number of global variant infections by the day of detection by variant origin continent, analogous to **F. (H)** The relationship between a country’s sequencing rate and the mean time to first global detection of a variant emerging in that country in metapopulation simulations (*N* = 160 for each variant *R*_*e*_). Lines correspond to LOESS fits by variant *R*_*e*._

Early detection of (variant) viruses such as potential zoonotic reassortant influenza viruses or highly genetically divergent SARS-CoV-2 variants maximises the time available to characterise the threat posed and design and implement potential interventions and mitigation strategies^1,6–8^. Hence, it is paramount for minimising potential public health impacts. To guide efforts toward improved preparedness for future respiratory virus threats, it is important to understand how the global landscape of genomic surveillance capabilities impacts the ability to swiftly identify new respiratory viruses and their variants. Furthermore, planning towards enhanced preparedness requires meaningful minimum sequencing targets as well as functional upper bounds for effective and efficient detection of new (variant) viruses^2,3,5,9–11^.

We aimed to quantify how the global distribution of clinical genomic surveillance infrastructure affects the global capacity to rapidly detect and characterize the spread of a novel respiratory virus (variant). First, we used a mathematical model to determine a target minimum global sequencing capacity that balances effective performance and efficient resource use. Then, we leveraged large-scale epidemic simulations to investigate how varyingly solidaristic global distributions of genomic surveillance infrastructure affect surveillance effectiveness. We used 2022 SARS-CoV-2 sequencing output as baseline, representing an empirical pandemic scenario with unprecedentedly high but highly unequally distributed levels of virus genomic sequencing.

## Results

### Global variation in pandemic-period detection capacity

To investigate how global variation in genomic surveillance capacity impacts the speed of new variant detection, we first investigated the performance of global genomic surveillance efforts for SARS-CoV-2 in 2022, representing an empirical baseline expectation for a potential future pandemic scenario. Sequencing output in 2022 was highly unequally distributed: country-specific sequencing rates estimated from submissions to GISAID^12^ ranged from <0.01 sequences per million people per week (S/M/wk) in some countries to >1000 S/M/wk in others (Fig. 1a). The median sequencing rate across European countries amounted to 64.3 S/M/wk, compared to 0.18 S/M/wk for countries in Africa. Similarly, the median time from sample collection to deposition in GISAID (henceforth, turnaround time) ranged across countries from less than a week to hundreds of days (Fig. 1b).

To understand how this variation impacts potential global detection capacity, we used a global metapopulation model, validated against GLEAM^13,14^ (Extended Data Fig. 2), to simulate hypothetical scenarios of global variant spread and subsequent detection. We performed 10,000 independent simulations for values of variant *R*_*e*_ ranging from 1.2 to 2. We assumed a distinct archetypal scenario of variant emergence, characterized by initial *R*_*e*_ and prevalence of wildtype virus, for each value of variant *R*_*e*_ (Extended Data Fig. 3). In each simulation, the country where the variant emerged was randomly selected based on a country population size-weighted probability. We then simulated the time to first variant detection for each metapopulation epidemic simulation, given empirical country-specific SARS-CoV-2 sequencing rates and turnaround times in 2022 (Fig. 1a,b).

Averaged across simulated variant *R*_*e*_ values, the mean time to first variant detection globally was 83.0 days (95% CI 18 – 194), with substantial variability especially at lower values of variant *R*_*e*_ (Fig. 1c). The simulated global number of variant infections by the day of first global detection varied widely (mean 632,899 infections, 95% CI 77 – 5,917,647), spanning up to five orders of magnitude for all values of variant *R*_*e*_ (Fig. 1d). In many simulations, new variants were first detected outside of their continent of origin, driven especially by variants first emerging in Africa (first detected outside origin continent in 75.0% of simulations), Asia (23.5%) and South America (19.1%) (Fig. 1e). This means that the variant would have frequently spread widely within and between continents prior to initial detection (consistent with, for example, the early spread and detection of the SARS-CoV-2 Omicron BA.1 variant^1^). The continent in which the variant first emerged strongly shaped the time to variant detection (Fig. 1f) and the number of global variant infections by the day of first detection (Fig. 1g), the latter ranging from a mean of 23,006 infections (95% CI 29 – 242,943) when emerging in Europe to 1,757,677 infections (95% CI 1028 – 13,369,527) in case of emergence in Africa across simulated values of variant *R*_*e*_. Differences in time to variant detection were strongly and highly nonlinearly associated with the sequencing capacity in the country of origin of the novel virus, with low sequencing rates being associated with longer times to variant detection (Fig. 1h).

### Operationalizing global health solidarity

Globally, there is a shared risk of the emergence of pandemic viruses or their variants. In contrast, the results above indicate the capacity to rapidly detect new (variant) viruses is profoundly asymmetrically distributed. In part, this imbalance reflects a deficit of global health solidarity. Solidarity as a principle specifically underlies institutionalized forms of sharing as a result of mutual dependence^15,16^; it gives guidance to human action in the face of interdependency related to the shared risks of communicable disease, and underlies the obligations reflected in the IHR^17,18^. Given that pandemic risk is globally shared, we sought to investigate how more or less solidaristic approaches to respiratory virus genomic surveillance could lead to varyingly effective and efficient outcomes for purpose of the global detection of novel (variant) respiratory viruses. To do so, we specifically considered national respiratory virus genomic surveillance capacity. We first sought to identify a minimum sequencing capacity at the national level that could serve as a target toward improving global capacity for rapid global (variant) virus detection. Ideally, this target would ensure timely information for public health action as well as efficient use of potentially limited resources. It would also need to be realistically attainable and sustainable in pandemic and inter-pandemic periods.

To identify a target minimum global sequencing capacity, we explored the relationship between sequencing rate, turnaround time, and time to variant detection in any single country in more detail. Representing a scenario of emergence of a potential future pandemic respiratory virus (variant), we simulated the emergence of a variant virus in the background of circulating wildtype virus and computed the expected time to variant detection based on binomial sampling for different sequencing rates. We then derived a new mathematical model characterising the relationship between sequencing rates and time to detection of the new virus variant. For a variant virus, introduced in a population at an initial frequency *f*_0_, where the change in variant proportion through time can be described by a logistic growth rate *s*, the time since variant introduction when the variant virus is expected to have been detected with confidence level 1-*q* when sequencing *n* samples per unit time is equal to (log[(*q*^-*s*/*n*^-1)/*f*_0_]+1)/*s* (Extended Data Fig. 4). This model is applicable to all respiratory viruses that can be described by SIR dynamics^19^, including SARS-CoV-2, seasonal influenza virus, respiratory syncytial virus, and potential future pandemic respiratory viruses.

### Benefits of increases in sequencing rate are rapidly diminishing

For all modelled scenarios of variant emergence (Extended Data Fig. 3), time to variant detection rapidly decreased as sequencing rate increased up to ∼10 S/M/wk while the benefits of increases in sequencing rate beyond 10 S/M/wk were much smaller (Fig. 2a, Extended Data Fig. 5a). In 2022, many high-income countries sequenced SARS-CoV-2 genomes at rates well in excess of 10 S/M/wk, whereas sequencing rates in many lower-and-middle-income countries were such that, in absolute terms, small increases would substantially speed up variant detection (Fig. 2a, Extended Data Fig. 5a). For example, in a country of 100 million people sequencing at the median 2022 SARS-CoV-2 sequencing rate in low-income countries (0.035 S/M/wk), increasing the sequencing rate by 1 S/M/wk would reduce the time to detection of a variant with *R*_*e*_ = 1.6 at 95% confidence by ∼28 days, given a wildtype prevalence of 0.5% and a wildtype *R*_*e*_ of 1.1 at time of variant emergence. In contrast, if the same country was sequencing at the 2022 median high-income country rate (58.9 S/M/wk), the reduction in time to detection resulting from the same 1 S/M/wk increase in sequencing rate would be only 3.5 hours (Fig. 2a, Extended Data Fig. 5b). The diminishing returns at rates characteristic of high-income countries are particularly prominent when looking at the relationship between sequencing rate and the expected number of variant infections by the day the variant has been detected (Fig. 2b, Extended Data Fig. 6a). Assuming a 14-day turnaround time, increasing the sequencing rate in a country sequencing at the median low-income country rate by 1 S/M/wk would reduce the expected number of variant infections by the time of detection with 95% confidence by ∼4.5 million infections for the scenario of variant emergence described above; in a country sequencing at the median high-income rate, the same 1 S/M/wk increase would only reduce the expected number of variant infections by the day of first detection by ∼60 infections (Fig. 2b, Extended Data Fig. 6b).

**Figure 2.**
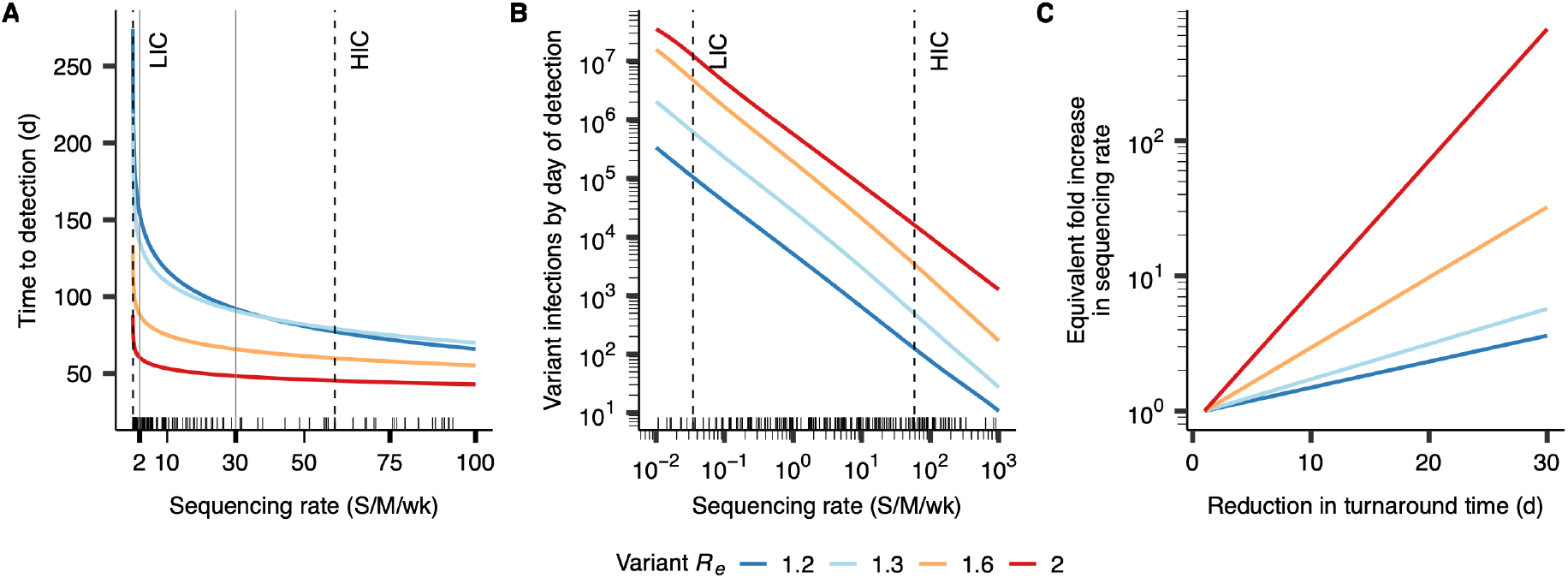
The dependence of time to variant detection on sequencing rate and turnaround time for a single country. **(A)** Relationship between sequencing rate and the number of days until the variant will have been detected with 95% confidence. The small black tick marks on the *x*-axes in this plot and in B show country-specific SARS-CoV-2 sequencing rates for 2022. Vertical dotted lines correspond to the median SARS-CoV-2 sequencing rates for high-income (HIC) and low-income (LIC) countries in 2022. In all panels, lines are coloured by values of variant *R*_e_, with a distinct scenario of variant emergence for each value of variant *R*_e_; sequencing turnaround time was assumed to be 14 days. **(B)** Relationship between sequencing rate and the expected number of variant infections by the day the variant will have been detected with 95% confidence. **(C)** Relationship between a reduction in turnaround time (in days) and the fold increase in sequencing rate that would be required to effect the same reduction in time to detection if turnaround time was kept constant.

In addition to sequencing rate, turnaround time is an essential component of effective genomic surveillance^2,5,20,21^. For reducing time to variant detection, any reduction in turnaround time is functionally equivalent to a fold increase in sequencing rate (Fig. 2c). Reductions in turnaround time are especially valuable for the detection of variant viruses that are highly transmissible. For example, for the archetypal variant with *R*_*e*_ = 2, a three-week reduction in turnaround time is equivalent to an 89.0-fold increase in sequencing rate (Fig. 2c). Hence, the benefits of increasing sequencing output should be carefully weighed against the gains from strengthening the ancillary infrastructure necessary for timely availability of sequencing results.

Using these results, we identified a possible target for a global minimum sequencing capacity. Given the identified relationship between sequencing rate, turnaround time, and time to detection, a sequencing capacity of 2 S/M/wk with a two-week turnaround time is a sensible potential global minimum target (Fig. 2a, vertical grey line). Its position at the elbow of the relationship between sequencing rate and time to detection (Fig. 2a) suggests that 2 S/M/wk is efficient, and its rapid variant detection even when a highly transmissible variant emerges in the background of high wildtype prevalence suggests that it results in strong performance. We chose a relatively low turnaround time of fourteen days given the vital importance of turnaround time in shaping time to detection. A sequencing rate of 2 S/M/wk corresponds to 0.18% of the maximum country-specific SARS-CoV-2 sequencing rate in 2022. If all countries sequencing at rates lower than 2 S/M/wk in 2022 were to attain this minimum capacity, the *de novo* generated sequencing capacity would represent 6.0% of global sequencing output in 2022. This suggests that in terms of raw sequencing capacity, the expansion necessary to effectuate the global minimum would be modest compared to empirical global sequencing output in a pandemic scenario.

### Global solidarity improves surveillance effectiveness and efficiency

To model the effect of more solidaristic global genomic surveillance, we re-simulated the global (variant) virus detection process given the global metapopulation epidemic simulations in a scenario where all countries possessed a global minimum capacity of at least 2 S/M/wk with 14-day turnaround time. Ensuring this global minimum sequencing capacity globally while keeping sequencing output unchanged for countries that already satisfied the minimum requirement in 2022 (henceforth, strategy A) reduced mean time to global variant detection by 26.0 days to 57.0 days (95% CI 17 – 119) relative to the simulated 2022 baseline (red bar, Fig. 3a). The mean number of global variant infections by the day of detection decreased from 632,899 infections (95% CI 77 – 5,917,647) to 31,485 infections (95% CI 67 – 235,057) (red bar, Fig. 3b), and the probability that the variant was first detected in its origin continent increased from 71.0% to 96.4% (red cross, Fig. 3c).

**Figure 3.**
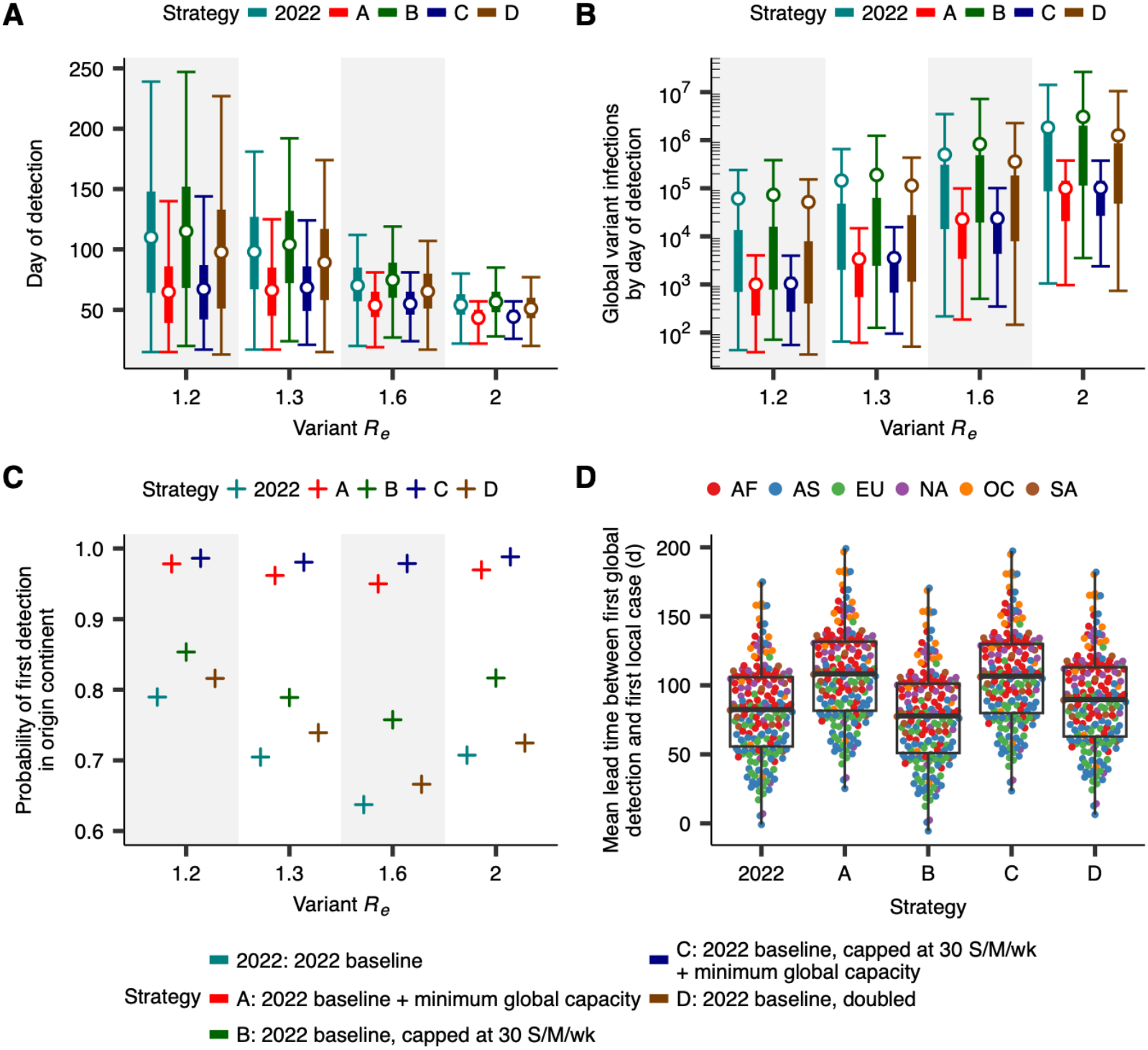
The time to first global detection of a new variant under varying global distributions of global respiratory virus genomic surveillance infrastructure. **(A)** Comparison of time to variant detection for different global strategies for the global distribution of genomic surveillance infrastructure. Each value of variant *R*_*e*_ corresponds to a distinct scenario of variant emergence (*N* = 10,000 for each). Thin and thick lines correspond to 95% and 50% CIs, respectively. Points correspond to means. **(B)** The cumulative number of global variant infections by the day of variant detection by strategy, analogous to A. **(C)** The probability that the variant is first detected in its origin continent, by strategy. **(D)** Comparison of the mean time between the first detection of the variant globally, and the first local within-country infection, by strategy, for individual countries, averaged across values of variant *R*_*e*_ (*N* = 195 for each strategy). Each point corresponds to a country, coloured by continent (AF: Africa, EU: Europe, OC: Oceania, AS: Asia, NA: North America, SA: South America). Boxplots show the median, first and third quartiles, and minimum and maximum values.

Since reductions in time to detection resulting from increases in sequencing rate beyond ∼10 S/M/wk (Fig. 2b) are limited, we further hypothesized that relative to the 2022 baseline, limiting sequencing rates to 30 S/M/wk would have little detrimental effect on time to variant detection (Fig. 2a, vertical grey line). In our simulations, setting a 30 S/M/wk upper limit in all countries relative to the 2022 baseline but no minimum requirement (henceforth, strategy B) left the expected time to first global variant detection (green bar, Fig. 3a) and the expected number of variant infections by the day of detection (green bar, Fig. 3b) largely unchanged: mean time to variant detection increased by only 4.6 days, from mean 83.0 days to 87.6 days (95% CI 22-202) (green bar, Fig. 3a), while global sequencing output was reduced by 67.0%.

To model a globally solidaristic approach to respiratory virus genomic surveillance, we combined the insights that establishing a global minimum sequencing capacity could strongly reduce time to variant detection whereas the reductions in time to detection beyond ∼10 S/M/wk rapidly diminish. Simulations indicated that in a hypothetical future pandemic scenario, ensuring a minimum global capacity of 2 S/M/wk, while also setting a 30 S/M/wk upper limit (henceforth, strategy C), could improve time to variant detection by weeks while still reducing sequencing output by 61.0% relative to the 2022 pandemic baseline (blue bar, Fig. 3a, Extended Data Fig. 7). This result suggests that achieving a global minimum surveillance capacity could allow for substantial improvements in the capacity for rapid global virus detection, even with substantially fewer total viruses sequenced globally. We also investigated a scenario of independent country-level expansion, where each country’s sequencing output increased proportional to its existing rate. Independently doubling each country’s 2022 sequencing output, (strategy D) would only reduce mean time to detection by 7.2 days (brown bar, Fig. 3a, Extended Data Fig. 7), suggesting that siloed expansion of individual countries’ sequencing output cannot replace a solidaristic global approach.

Initial detection is a necessary starting point for responses to potential novel threats. However, additional information beyond simple detection is often necessary to characterize the public health risk that a (variant) virus poses. For example, the SARS-CoV-2 Alpha variant was first detected in the UK in a sample collected on 20 September 2020, likely within days of its initial emergence^22^. However, it was not until December 2020 that epidemiological evidence of the variant’s transmission advantage relative to pre-existing viruses began to accumulate^22,23^. To that end, we also investigated how a more solidaristic global distribution of sequencing output would affect the time elapsed until the variant would have been estimated to account for a substantial proportion of circulating virus, suggestive of a potential transmission advantage. In our simulations, the time until estimated variant frequencies, in at least one country, provided evidence with 95% confidence that the variant had reached 1% circulating frequency, decreased from 117.7 days (95% CI 42-252) for the 2022 baseline to 103.8 days (95% CI 42-210) for strategy C (Extended Data Fig. 8a). Correspondingly, the mean number of global infections by that day decreased from 2,072,633 (95% CI 4,896-20,293,166) to 205,311 (95% CI 4,873-1,428,529) (Extended Data Fig. 8b). In contrast, capping sequencing rates at 30 S/M/wk (strategy B) increased the mean time until the variant was established to have reached 1% circulating frequency somewhere globally by only 0.5 days relative to the 2022 baseline (Extended Data Fig. 8a). Mathematical models indicate that a sequencing capacity of 2 S/M/wk would ensure robust ascertainment of variant prevalence (Extended Data Fig. 9).

### Solidaristic approaches improve opportunities for mitigation

To investigate how establishing a global minimum capacity could affect public health preparedness, we computed the mean lead time between first global detection and the first local case for all countries under the different strategies. As the first global detection of a (variant) virus represents a potential starting point for the design and implementation of local public health responses, this lead time provides a measure for individual countries of the time horizon for public health measures that aim to mitigate potential impacts. In all countries, the lead time would increase under more solidaristic distributions of global sequencing infrastructure (Fig. 3d), potentially allowing for more time to implement public health measures in preparation for variant outbreaks or nascent pandemics. For example, for the archetypal variant virus with *R*_*e*_ = 1.6, the mean time between first global detection and arrival in the United States was -8.5 days under the 2022 baseline, suggesting that on average, the variant would already be present in the United States by the time it was first detected globally. Under the solidaristic strategy C, the public health lead time in the US increased by two weeks to +6.6 days. The increases in lead time were stronger for lower values of *R*_*e*_; for example, for the archetypal variant with *R*_*e*_ = 1.3, the mean lead time increased from +114 to +144 days in Rwanda, +82 to +111 days in Kazakhstan, +47 to +76 days in Indonesia, and +21 to +51 days in the United Kingdom, for strategy C relative to the 2022 baseline.

## Discussion

Our results indicate that operationalizing global health solidarity in respiratory virus genomic surveillance could strongly improve preparedness for potential future respiratory virus threats. Relative to siloed surveillance efforts, where countries’ policies are strongly domestically focused, pursuing sustainable global capacity could substantially reduce the time to first global detection of variant viruses and the time until variant viruses are found to exhibit signatures of rapid spread. Initial detection and sequencing is a necessary first step in assessing and responding to the threat posed by novel viruses and underlies the design and deployment of countermeasures such as vaccines and therapeutics^7,24^. As such, earlier warning of potential threats could substantially improve the time horizon for global and local public health measures that aim to mitigate viral threats’ potential impacts, and solidaristic approaches to global genomic surveillance could improve outbreak preparedness and response for all countries globally.

Our results suggest that only a small fraction of pandemic-period sequencing output, in the right places, could transform the global capacity to rapidly detect novel threats. This fact, combined with the fact that a country can only detect a virus that emerged elsewhere once it is already present locally, suggests that to improve outbreak preparedness, siloed expansion of surveillance capacity in countries that already possess strong capacity cannot replace solidaristic global investment, and that solidaristic approaches to genomic surveillance could yield greater public health benefit even to countries that already possess strong surveillance infrastructure locally. Our analyses are primarily focused on detecting novel (variant) viruses and tracking their spread. Hence, our arguments weighing the enhancement of local surveillance capacity against the development of basic global capacity do not consider ancillary benefits of high-intensity genomic surveillance in high-income settings such as characterization of local transmission dynamics. However, we stress the fundamental immediate importance of basic global capacity for initial detection for all countries, such as in the context of vaccine development and deployment, where speed-ups of a few weeks could have substantial public health impacts globally^25^.

In our model we assumed representative sampling in the genomic surveillance process, including the ready availability and access to diagnostic tools, which does not always hold in reality^26,27^. As the departure from this assumption is especially strong in resource-constrained settings^20,26^, the reported reductions in time to variant detection resulting from the establishment of a global minimum sequencing capacity are likely underestimates. Furthermore, our model does not model the spatial distribution of the minimum capacity in the country. The spatial distribution of sequencing capacity and the structure of sample referral networks interplays with turnaround time to shape surveillance performance and affects the optimal country-level implementation of surveillance networks. Our model is not applicable to (variant) viruses with no or a detrimental effect on transmissibility such as those that only result in increased disease severity or reduced sensitivity of diagnostics. Importantly, our results are robust to biases in the estimates of turnaround time resulting from delays in sequence deposition in GISAID^28,29^ (Extended Data Fig. 10a) and deviations from the assumed global mobility rates (Extended Data Fig. 10b).

While solidarity is a helpful principle to guide policy in the case of an interdependence of risks given a particular context, reaching health equity or health justice will require overcoming substantial challenges^30–35^. Even with the capacity to detect new viruses sooner, the capacity to respond is also distributed asymmetrically. Even if the proposed models help undergird global health solidarity, the benefits of more rapidly available vaccines or better-matched vaccine updates due to timelier detection will only extend to countries with access to these benefits^30–32,36^. Furthermore, rapidly detecting and sharing information concerning new (variant) viruses must not paradoxically disadvantage countries that do so. Open sharing of pathogen genomic data must operate within a system of fair access and benefit-sharing to achieve its intended public health purpose without exacerbating global health inequity^33,36,37^. Despite these challenges, in many countries there is a desire for enhanced genomic surveillance capacity to inform local public health responses^21,27,29,38,39^.

Given the relationship between turnaround time and sequencing rate, we proposed a sequencing capacity of 2 S/M/wk with 14-day turnaround time as a target. We note that given this capacity, the optimal sequencing rate and its balance with turnaround time depends on the characteristics of the pathogen, the epidemiological background in which the variant were to emerge, and the required timeliness of sequencing data for public health action; for example, when levels of wildtype respiratory virus circulation are very low, relatively fewer sequences might yield a better balance between surveillance performance and resource use.

The proposed target aims to balance the resources necessary for surveillance in periods of seasonal circulation of respiratory pathogens with the capacity to rapidly detect and scale up capacity during potential pandemic scenarios. While our study is focused on respiratory viruses and their variants, leveraging the infrastructure associated with the proposed target for surveillance of non-respiratory pathogens would yield further benefits. Our results underscore the importance of turnaround time in shaping the effectiveness and public health utility of surveillance efforts^20,21^, and particularly its balance with sequencing rate. Relieving barriers to attaining low turnaround times in resource-limited settings, such as the availability of reagents, is key to realizing the potential benefits of global surveillance capacity^20,40^.

While our modelling results provide a principled target that balances resource use and performance, the optimal design, including the balance of sequencing rate and turnaround time, will likely differ from country to country, depending on local constraints and priorities. Our study provides quantitative evidence of how solidaristic approaches provide a rational basis for improving global surveillance performance, but implementation requires addressing challenges related to infrastructure, personnel, and funding that currently form barriers to the implementation of genomic capacity in under-resourced settings^29,40–42^. To achieve the long-term advancement of global genomics capacity in these settings, coherent capacity-building is necessary, and that requires sustainable, diversified financing which minimizes dependency on single funding source while aligning well with national needs^40^.

The COVID-19 led to an unprecedented expansion of sequencing capacity globally. Some of the most consequential gains were made in resource-limited settings^20,21^, and it essential that such gains are maintained and where necessary expanded to maximize preparedness for future threats. Our results suggest that a global minimum respiratory virus sequencing capacity offers a path toward improved responses to respiratory virus threats, even for countries with existing strong national surveillance capacities. For these countries, supporting a global minimum sequencing capacity could yield benefits in preparation for and during potential future outbreak scenarios that are not attainable through siloed focuses on local capacity. Our study shows how a global outlook on pandemic preparedness is essential to improve both global and local public health. To improve outbreak preparedness, there is no substitute for global solidarity; it offers a path toward better responses to respiratory virus threats that would be mutually beneficial to all WHO Member States.

## Methods

### Operationalizing global health solidarity

Genomic surveillance capacity has become a core component of preparedness and response to global disease outbreaks. In accordance with Article 5 and annex 1 of the International Health Regulations (IHR), Member States of the World Health Organization (WHO) must ensure national surveillance capacity. The WHO can assist Member States to develop, strengthen and maintain surveillance capacities as needed (Article 5(4) IHR)^4^. Underlying this obligation is solidarity; a principle that gives guidance to human action in the face of interdependency because of the shared risks of communicable disease^17,18^.

Solidarity connotes a sense of commitment to help similar others in need^43^. More specifically solidarity – quite apart from charity or other forms of ‘helping out’ and sharing – as a principle underlies institutionalized forms of sharing as a result of mutual dependence^15,16^. Solidarity is relational, meaning that its institutional scope is determined by societal bounds of whom we feel solidary towards^44,45^. In the field of health this plays out in national schemes of health insurance, redistribution, planning and rationing to ensure access to medicines and services^46,47^.

The relational aspect of solidarity has made it difficult globally to determine what exactly is owed in interstate and global health interactions. In this regard it has been argued that in global health, solidarity needs to be based on ‘similarity in a *specific context’*^43^. In line with this presupposition, we operationalize global health solidarity here specifically considering national capacity for genomic sequencing, by modelling for the most effective and efficient global distribution to the extent that we globally face the shared risk of unforeseeable pandemic respiratory virus occurrence. Operationalizing global health solidarity through modelling the global distribution of genomic sequencing capacity in this regard can guide WHO efforts to the implementation of Article 5 IHR, and to rationally invest in genomic sequencing capacity where needed to safeguard global pandemic preparedness as risk-sharing among WHO’s Member States.

### Sequence metadata analysis

We downloaded metadata corresponding to all SARS-CoV-2 genomes in the GISAID^12^ database with collection date from January 1^st^ to December 31^st^ 2022 and submission date before July 1^st^ 2023 (*n* = 6,914,601). For each country with at least one sequence in the dataset, we computed the weekly sequencing rate by dividing the number of viruses sampled in that country by 52 and the country’s population size in millions, yielding a sequencing rate in units of sequences per million people per week (S/M/wk). Population sizes for July 1^st^ 2022 were extracted from the United Nations World Population Prospects 2022 (https://population.un.org/wpp/Download/Standard/MostUsed/). For each sequence, we computed the turnaround time from the number of days between the sample collection and submission day in GISAID. We extracted countries’ *per capita* gross domestic product (GDP) for 2022, or the most recent year before 2022 if data for 2022 was unavailable, from the World Bank (https://data.worldbank.org/indicator/NY.GDP.PCAP.CD (last updated 2023/10/26)). We extracted income classifications for each country for fiscal year 2024 from the World Bank (https://datahelpdesk.worldbank.org/knowledgebase/articles/906519-world-bank-country-and-lending-groups).

### Variant epidemic simulations

In all analyses, we assumed that a variant virus emerges in the context of circulating wildtype virus. In our simulations, both variant and wildtype epidemiological dynamics are described by a susceptible-infected-recovered (SIR) compartmental model with infectious period 1/*γ* equal to 5 days for both viruses, with no interactions between genotypes. We simulated variant epidemics under a range of values of variant *R*_*e*_ at time of introduction (variant *R*_*e*_ = 1.2, 1.3, 1.6, and 2). In the main text, we assumed a different scenario of variant emergence for each value of variant *R*_*e*_, characterized by a wildtype (wt) *R*_*e*_ at time of variant introduction and a wildtype prevalence at time of variant introduction (variant *R*_*e*_ = 1.2: wt *R*_*e*_ = 1, wt prevalence = 0.1%; variant *R*_*e*_ = 1.3: wt *R*_*e*_ = 1.05, wt prevalence = 0.2%; variant *R*_*e*_ = 1.6: wt *R*_*e*_ = 1.1, wt prevalence = 0.5%; variant *R*_*e*_ = 2: wt *R*_*e*_ = 1, wt prevalence = 2%). These scenarios were chosen such that circulation dynamics of wildtype and variant were comparable (e.g. the emergence of a highly transmissible variant in the background of high wildtype prevalence). In the Extended Data Figures, we show the same analyses for all combinations of variant *R*_*e*_ and scenario of variant emergence (e.g. a variant with *R*_*e*_ = 2 with wildtype dynamics corresponding to the scenario for variant *R*_*e*_ = 1.2 (wt *R*_*e*_ = 1, wt prevalence = 0.1%)). Epidemic dynamics for each scenario in the main text are shown in Extended Data Fig. 3. While we report results based on variant *R*_*e*_, we note that results are primarily dependent on the logistic growth rate of the variant proportion (see section ‘*Mathematical model*’ below). In this context, the scenario of variant *R*_*e*_ = 2 with wt *R*_*e*_ = 1 is, for example, functionally equivalent to a scenario with variant *R*_*e*_ = 2.5 and wt *R*_*e*_ = 1.5. Similarly, the first scenario of variant *R*_*e*_ = 1.2 and wt *R*_*e*_ = 1 is functionally equivalent to a scenario of variant *R*_*e*_ = 1.6 and wt *R*_*e*_ = 1.3, approximating a scenario of emergence of a A/H1N1pdm09 pandemic-like virus early in a seasonal influenza virus epidemic.

### Metapopulation model

We used a metapopulation model that couples local SIR dynamics within each index country with global migration to simulate the global spread of a variant. Given a rate of movement *w*_*nm*_ from population *m* to *n*, the expected number of variant-infected (*I*_*n*_) and variant-susceptible (*S*_*n*_) people in population *n* with population size *N*_*n*_ for a variant with transmission rate *ϕ* and recovery rate *γ*, is described by

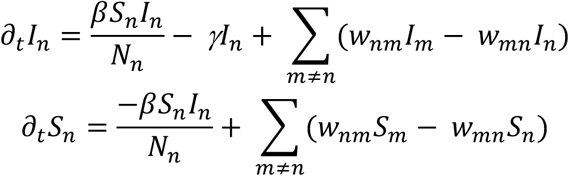

This model is the basis of the model used by Brockmann et al.^48^ to fit empirical arrival times for multiple respiratory viruses to global air transportation data. We used the estimated pairwise number of trips between all countries from the Global Transnational Mobility (GTM)^49^ to inform *w*_*nm*_. This dataset combines a tourism dataset from the World Tourism Organization and an origin-final destination dataset corresponding to global air travel data. Previous work has validated the GTM against the world airline network^50^, which Brockmann et al. ^48^ showed to strongly reproduce observed dynamics of global pathogen spread. Specifically, for any two countries *n* and *m* we computed *w*_*nm*_ by dividing the number of trips from country *m* to *n* in the year 2016 by the population size of country *m* and by 365. For each value of variant *R*_*e*_, we performed 10,000 independent simulations of the metapopulation model, assuming that the probability a variant virus would emerge in a particular country is proportional to the country’s relative population size (simulations initialized in Africa: *n* = 1793; Asia: *n* = 5946, Europe: *n* = 934; North America: *n* = 739; Oceania: *n* = 54; South America: *n* = 534). We integrated the model forward in time at a daily timescale using a tau-leap algorithm, which also furnishes the epidemic dynamics and global spread with stochasticity. Each simulation was initialized with an infected population of 10 individuals.

We validated the metapopulation model by comparing arrival times against those that were independently estimated using GLEAM^14^, a separate metapopulation model that incorporates commuting but which relies on different underlying data. Given an epidemic origin location, we simulated 10 epidemic instances using the metapopulation model, each initialized with 10 infected individuals, and we simulated 10 instances using GLEAM, where we implemented the same SIR model. In the GLEAM simulations, we assumed 100% of airline traffic, no seasonality, and a gravity commuting model with 8 hours spent at the commuting destinations. For each country, we computed the first day on which median cumulative incidence across simulations exceeded 0.01 per 1000 individuals for both model implementations. We performed these simulations for ten countries (Cameroon, Ecuador, France, Jamaica, Malaysia, Mali, Nepal, Nicaragua, Oman, Uzbekistan) with the GLEAM model initialized in each country’s capital city. For all ten origin locations we find a strong concordance between arrival times *(r* = 0.89 overall) estimated using the metapopulation model and GLEAM (Extended Data Fig. 2). This provides support for the use of the metapopulation model.

### Global genomic surveillance simulations

We performed the genomic surveillance simulations using empirical turnaround times and sampling rates for each country, using data for 2022. For each sequence in GISAID, we computed the time *T* between the sample’s collection date and submission date. For each country *c*, the turnaround-time specific sequencing rate in unit of sequences per day *n*_*x,c*_, for each value of turnaround time *x* in days, was equal to the country’s total sequencing rate in sequences per day multiplied by the proportion of sequences from that country with *T* = *x*.

For each country in each simulation, starting from the first day on which the number of new variant infections exceeded 10 onwards, we deterministically simulated the wildtype epidemic dynamics. For each value of variant *R*_*e*_, we assumed a scenario of variant emergence (characterized by a wildtype prevalence and wildtype *R*_*e*_) as described above in the main text. In the Extended Data Figures, we show the same analyses for all combinations of variant *R*_*e*_ and scenario of variant emergence. Until the first day on which the number of variant infections exceeded 10, wildtype incidence was assumed to be equal to wildtype incidence on the first day of the simulated wildtype epidemic, to account for the stochasticity observed when the number of infections was small and the potential for stochastic variant extinction.

Using the simulated variant and wildtype incidence on each day, we computed the variant proportion through time *f(t)*. For each country *c*, on each day *t*, we used the simulated country-specific variant proportion *f*_*c*_(*t*) to simulate genomic surveillance: For each value of turnaround time *x*, we assumed that total sample count *ñ*_*x,c*_ *∼* Poisson(*n*_*x,c*_) and simulated the total number of variant samples 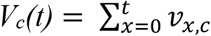, with *v*_*xc*_∼ Binomial(*ñ*_*x,c*_, *f*_*c*_(*t-x*)). In each of 10,000 replicate simulations, and for each strategy for the global distribution of surveillance infrastructure (see next section), we computed the detection day as the first day *t* on which *V*_*c*_*(t)* was at least one in at least one country *c*. We defined the detection country as the first country for which this held.

To investigate the time until the variant could be said to account for a substantial proportion of circulating virus in at least one country, we used the simulated weekly sequence counts to compute, for each country, if there was any week in the past in which the variant accounted for at least a proportion *π* of all samples collected that week with 95% confidence given a one-tailed binomial test for proportions. We performed this analysis on a weekly basis for each country, and the day on which the *p-*value for this binomial test declined below 0.05 in at least country, for any week in the past, was defined as the day the variant was established to account for a substantial proportion of circulating virus in at least one country globally. We chose *π* to be 1% for all countries with a population of 100 million individuals or fewer. We ensured a more flexible threshold for countries with a population larger than 100 million. For these countries, the threshold decreased proportionally as the population size increased, e.g. using a threshold of 0.5% for a population of 200 million individuals and a threshold of 0.1% in a population of 1 billion individuals.

### Global surveillance strategies

We investigated five strategies for the global distribution of sequencing infrastructure:

Strategy 2022: the 2022 baseline. For each country, turnaround time-specific sequencing rates were extracted from GISAID metadata.

Strategy A: the 2022 baseline + a global minimum sequencing capacity of 2 S/M/wk with 14-day turnaround time in each country. If a country already satisfied this requirement (i.e., the sum of turnaround time-specific sequencing rates with turnaround time ≤14 days was equal to or greater than 2 S/M/wk), its sequencing rates were unchanged relative to the 2022 baseline. If a country satisfied the sequencing rate across all values of turnaround time, but not within the required two-week turnaround time, the deficit in S/M/wk in the sum of turnaround time-specific sequencing rates with turnaround time ≤14 days was uniformly removed from the sequencing rates exceeding 14 days and added to the sequencing rate corresponding to a turnaround time of 14 days. Hence, in this scenario, total sequencing output remained unchanged, and the minimum sequencing capacity was attained by reducing turnaround time. If a country did not satisfy the minimum sequencing rate at all, all sequencing output corresponding to a sequencing rate >14 days was set to a turnaround time of 14 days. The remaining deficit in S/M/wk in the sum of turnaround time-specific sequencing rates with turnaround time ≤14 days was added to the sequencing rate corresponding to a turnaround time of 14 days.

Strategy B: Equivalent to the 2022 baseline, but individual countries’ sequencing output capped at 30 S/M/wk. Countries that sequenced at rates exceeding 30 S/M/wk had their sequencing output capped by dividing sequencing rate uniformly across all values of turnaround time such that total output across all values of turnaround time was equal to 30 S/M/wk.

Strategy C: A combination of strategies A and B. In countries that, after capping according to strategy B, did not satisfy the minimum sequencing rate of 2 S/M/wk with 14-day turnaround time, this minimum was ensured analogous to Strategy A.

Strategy D: The 2022 baseline, doubled. In each country, the sequencing rate in 2022 was doubled across all values of turnaround time. Hence, the absolute increase in sequencing output was greater in countries that had a higher baseline sequencing rate.

### Mathematical model

For the single-country analyses presented in Figure 2, we assumed a population of 100 million and turnaround time of two weeks. We deterministically simulated variant and wildtype epidemics, starting with one variant-infected individual, and computed the variant proportion *f(t)* through time. For each sequencing rate and given *f(t)*, we computed the expected day of detection with 95% confidence as the day on which the probability that zero wildtype sequences would have been binomially sampled up to and including that day declined below 0.05. On each day, the total number of samples to sequence was assumed to be a Poisson-valued random variable with rate given by the sequencing rate. For each sequencing rate, the day of detection was computed as the median across 100 replicates. To compute the equivalent fold increase in sequencing rate for each reduction in turnaround time, we computed the slope of a linear model that relates the logarithm of the sequencing rate to the simulated day of detection for 1 < *n* < 100 S/M/wk. In Extended Data Fig. 5 and 6 we show the analyses of time to detection for all combinations of variant *R*_*e*_ and scenario of variant emergence (e.g. a variant with *R*_*e*_ = 2 with wildtype dynamics corresponding to the scenario for variant *R*_*e*_ = 1.2 (wt *R*_*e*_ = 1, wt prevalence = 0.1%)).

To mathematically model time to variant detection, we assumed that the variant frequency follows a logistic growth function, where the proportion *f(t)* of all new infections at time *t* that is attributable to the variant follows:

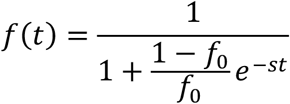

Here, *s* is the logistic growth rate that defines the speed at which the variant displaces the wildtype and *f*_0_ represents the initial variant frequency. The dynamics of logistic growth of variant proportion characterized the sequential replacement of variants during the COVID-19 pandemic. Assuming no interactions between genotypes, the value of *s* is equal to the difference of variant and wildtype exponential growth rates^23^. In reality, *s* is governed by factors such as pre-existing immunity in the population and differences in epidemiological characteristics of variant and wildtype such as their generation interval. Nevertheless, the derived relationship relies solely on the value of *s*, and hence is agnostic to the precise epidemiological characteristics of wildtype and variant. Given these dynamics, we derived a relationship between the number of viruses to sequence per unit time *n* and the expected time until the variant is detected. Beginning with the binomial probability that variant is detected at or before time step τ:

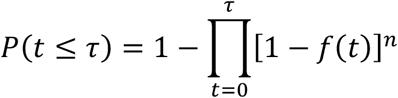

we derived an expression for τ:

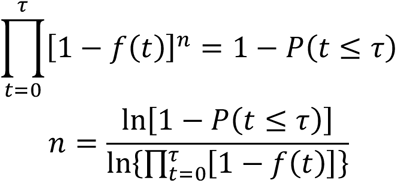

Using the Volterra product integral:

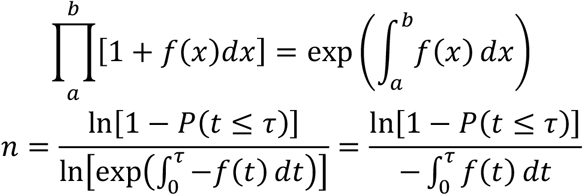

Integrating *f*(*t*):

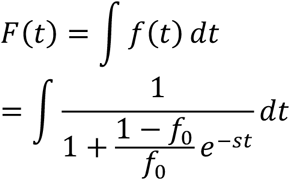

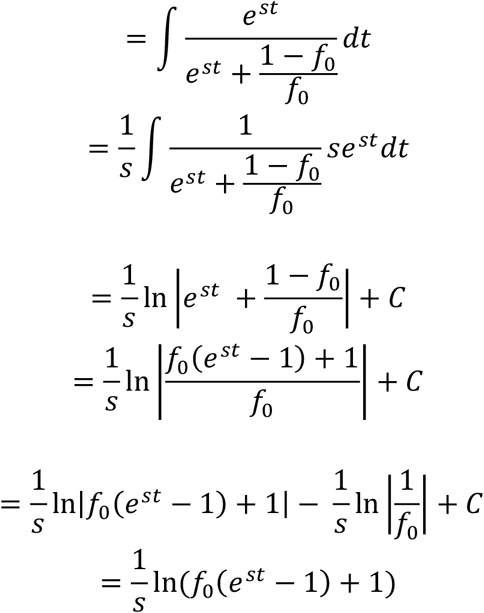

We can then rewrite:

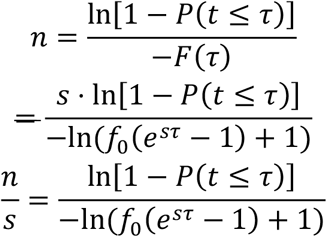

Let *q* = 1 − *P*(*t* ≤ τ) which is the probability that the variant will *not* be detected before or during time step τ.

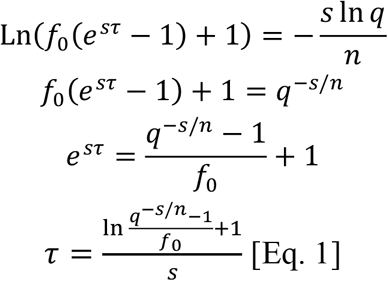

This equation yields, given *s, n, f*_0_, and *q*, the day τ on which the variant will have been detected at least once with confidence level 1 – *q*. This equation is valid when the timescales of detection are smaller than the timescales at which the logistic growth dynamics do not hold. For example, in extreme scenarios of a very high wildtype *R*_*e*_, a small variant transmission advantage and a low sequencing rate, the timescale of variant detection is beyond that of depletion of the susceptible population and the assumptions of the equation are not satisfied. We compared the predicted time to detection at 95% confidence for sequencing rates *n* ranging from 0.1 to 1000 S/M/wk as computed using epidemic simulations (Fig. 2a in main text) to predicted time to detection using only Eq. 1. In computing time to detection using the equation, we used the empirical value of *f*_0_ from the epidemic simulations as input, with *q* = 0.05. We used the theoretical value of *s*, computed as (variant *R*_*e*_ - wildtype *R*_*e*_) / 5. We performed this simulation for all four scenarios of variant emergence (each corresponding to a different initial wildtype *R*_*e*_ and wildtype prevalence) and all four values of variant *R*_*e*_. As seen in Extended Data Fig. 4, there was high correspondence between the time to detection from the explicit epidemic simulations and Eq. 1 when the variant *R*_*e*_ was high and/or wildtype prevalence was low. In contrast, when variant *R*_*e*_ was low and wildtype prevalence was high, susceptible depletion would occur before the timescale at which the variant would be detected, and the time to detection as predicted using the equation would deviate from the simulated time to detection. We note that, for combinations of initial variant proportion and variant proportion logistic growth rate not explicitly discussed in this study, the mathematical model can be used to compute the expected time to variant detection.

### Variant prevalence estimation

In addition to variant detection, we investigated the relationship between sequencing rates and the accuracy with which the spread dynamics of the variant can be tracked following its detection. Specifically, we investigated the accuracy with which the weekly proportion of new infections that is attributable to the variant can be estimated, and how this accuracy depends on sequencing rate. Mathematically, assuming a small, finite population *N* was infected at prevalence ρ and samples were collected from fraction *s* of infected individuals during each week, the potential (finite) number of samples that could be sampled from for sequencing is *N*ρ*s*.

Suppose the true circulating proportion is *p* and *n* (i.e. *n* < *N*ρ*s*) number of samples were sequenced, the number of variant sequences (*X*) follows a hypergeometric distribution with mean and variance:

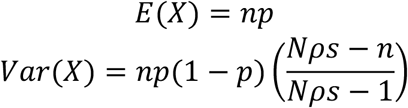

The variance of the variant proportion 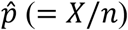 showing up in the sequences is:

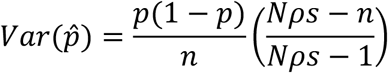

By Central Limit Theorem,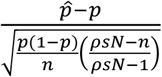 follows an approximate Normal distribution.

As such, at 95% (α = 5%) confidence, the error (*ϵ*) around the true variant proportion is:

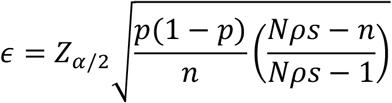

For sequencing rate of *r* sequences per million persons per week (hence 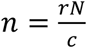 where *c* = 10^6^):

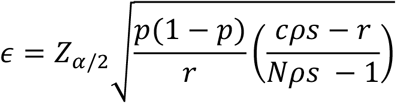

If *N*ρ*s* is sufficiently large (i.e. 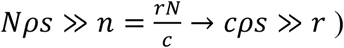,

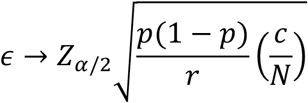

In Extended Data Fig. 9, we visualized the relationship between sequencing rate and the error in the estimated variant proportion, for different population sizes, true variant proportions, and values of α.

### Sensitivity analyses

In our analyses, we defined a sequence’s turnaround time as the time between the sequence’s collection date and its submission date on GISAID. This represents the most accurate measure of turnaround time available and has been used in previous analyses of global sequencing output^5^. Nevertheless, a potential issue with this definition of turnaround time is a lag between acquiring the sequence and its submission to GISAID, which is not reflected in these estimates^28^; in some cases, sequence analysis might have been performed but the sequence would only later be deposited in GISAID. To establish the sensitivity of our global simulation results to such delays in upload to GISAID, we re-simulated our global metapopulation genomic surveillance simulations, where we assumed that the day the sequence was acquired was somewhere between the sample’s collection date and date of submission to GISAID. Specifically, for each sequence, we computed the modified turnaround time as *ϕ*(*t*_*submission*_ – *t*_*collection*_), for 0 < *ϕ* < 1. We re-simulated the genomic surveillance simulation results as presented in Figure 3 for *ϕ* = 0.25, 0.5. Varying *ϕ* modifies the (country-specific) turnaround time-specific sequencing rates used in the global genomic surveillance system. Results for different values of *ϕ* are presented in Extended Data Fig. 10a. For all values of *ϕ* tested, we find that the conclusion holds that more solidaristic strategies for the global distribution of respiratory virus surveillance infrastructure (strategies A and C) offer strongly reduced time to variant detection. Hence, our results are robust to biases resulting from deviations from the assumption that the submission date represents the date on which the sample is available. Importantly, the observed consistency between a country’s sequencing rate and its median turnaround time (Spearman’s *ρ* = -0.60, *P* = 5.1× 10^−21^) suggests that a country’s distribution of turnaround times as computed from GISAID yields a representative picture of a country’s true capacity to rapidly sequence a virus after sample collection.

In our model, we estimated the mobility rate *w*_*nm*_ for countries *m* and *n* by dividing the number of trips from *m* to *n* in 2016 in the GTM by the population in *m*. This assumes that all members of the population participate in disease-relevant spread. In reality, this will not be the case. However, for the results of our study, these differences are likely to be of little consequence. Specifically, a lower effective mobility rate would further increase the reduction in time to detection that would result from the establishment of minimum sequencing infrastructure globally, as the time until a variant that emerges in a low-sequencing rate environment is exported to a high-sequencing rate environment would increase. We explicitly investigated the potential effects of misspecification of the mobility matrix on our results by multiplying and dividing the mobility rate matrix by three, representing substantially increased and reduce spread, respectively (Extended Data Fig. 10b). A reduced rate would further increase the gains to be effected by the establishment of sequencing infrastructure globally. Even if the mobility rate was increased three-fold, the strategies with increased solidarity in the global distribution of genomic surveillance infrastructure yield strongly improved performance compared to the 2022 baseline. Hence, our results are robust to specifics of the mobility dynamics.

Our analyses wholly rely on GISAID data to inform the global landscape of respiratory virus genomic surveillance infrastructure. In some countries, incomplete or absent deposition of sequence data in GISAID may result in sequencing rates computed from GISAID data being unreliable. For example, the zero-covid policy in China that was in place for parts of 2022, combined with a relatively small number of sequences in GISAID, suggest that submission rates to GISAID may not accurately represent China’s true genomic surveillance capacity^51^. We tested the possible implications of such biases on our results by comparing the results if all epidemic simulations with new variant viruses originating in China were removed. When China was removed from the set of possible epidemic origin locations, the conclusions regarding the performance of the different strategies for the global distribution of genomic surveillance infrastructure remained unchanged. The representativeness of GISAID data is further supported by the extremely strong correlation between sequence output and GDP (Spearman’s *ρ* = 0.79, *P* = 6.3×10^−41^).

## Data Availability

Custom code and data used to generate the results in this study is publicly available at https://github.com/AMC-LAEB/genomic_surveillance_solidarity. Raw global epidemic simulation output is available at https://zenodo.org/records/10051237.

## Data availability

Data on global population sizes are available from the United Nations World Population Prospects 2022 (https://population.un.org/wpp/Download/Standard/MostUsed/). Data on country GDP (https://data.worldbank.org/indicator/NY.GDP.PCAP.CD) and income classification (https://datahelpdesk.worldbank.org/knowledgebase/articles/906519-world-bank-country-and-lending-groups) is available from the World Bank. The Global Transnational Mobility Dataset is available from the Global Mobilities Project (https://migrationpolicycentre.eu/globalmobilities/dataset/). Metadata on global SARS-CoV-2 and seasonal influenza virus sequencing rates were extracted from GISAID (www.gisaid.org). Raw global epidemic simulation output is available at https://zenodo.org/records/10051237.

## Code availability

Custom code and data used to generate the results in this study is publicly available at https://github.com/AMC-LAEB/genomic_surveillance_solidarity.

---

**Extended Data Fig. 1.**
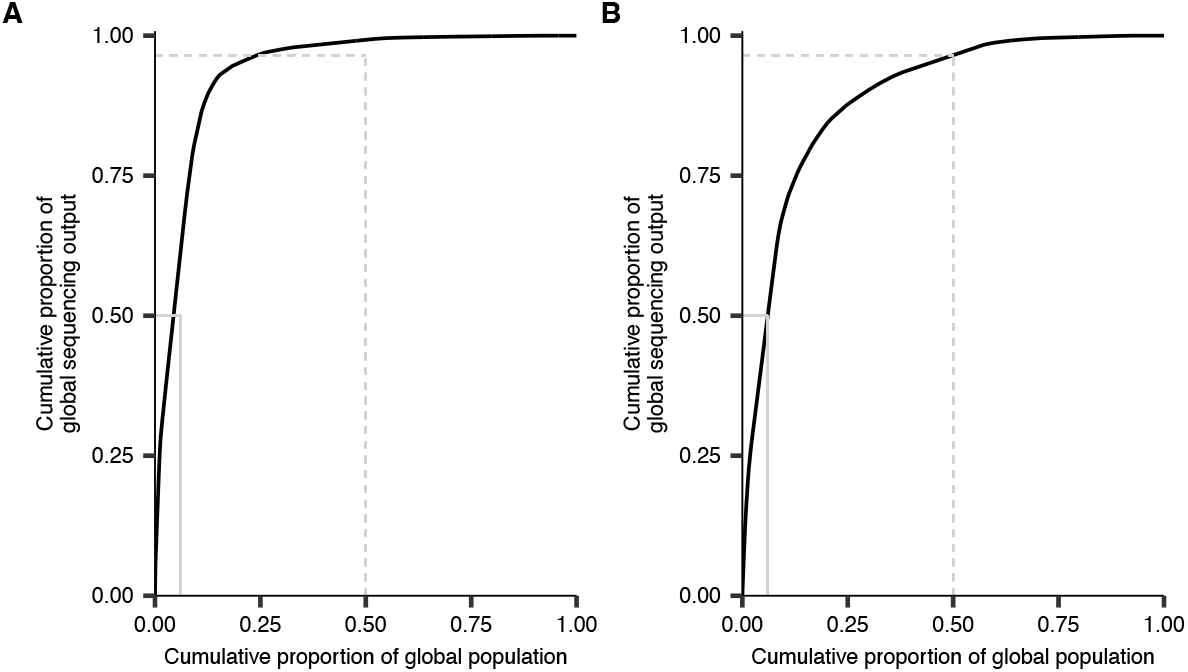
The global distribution of sequencing output. **(A)** The cumulative proportion of the global population that accounts for a cumulative proportion of global sequence output for SARS-CoV-2. Data on sequencing output corresponds to SARS-CoV-2 sequences in GISAID with collection date in 2022. Solid grey lines show the smallest proportion of the population that accounts for 50% of sequencing output. Dashed grey lines show the smallest proportion of sequencing output that is accounted for by 50% of the global population. **(B)** The cumulative proportion of the global population that accounts for a cumulative proportion of global seasonal influenza sequence output. Data on sequencing output corresponds to seasonal influenza sequences collected from humans in GISAID with collection date in 2018. Solid grey lines show the smallest proportion of the population that accounts for 50% of sequencing output. Dashed grey lines show the smallest proportion of sequencing output that is accounted for by 50% of the global population.

**Extended Data Fig. 2.**
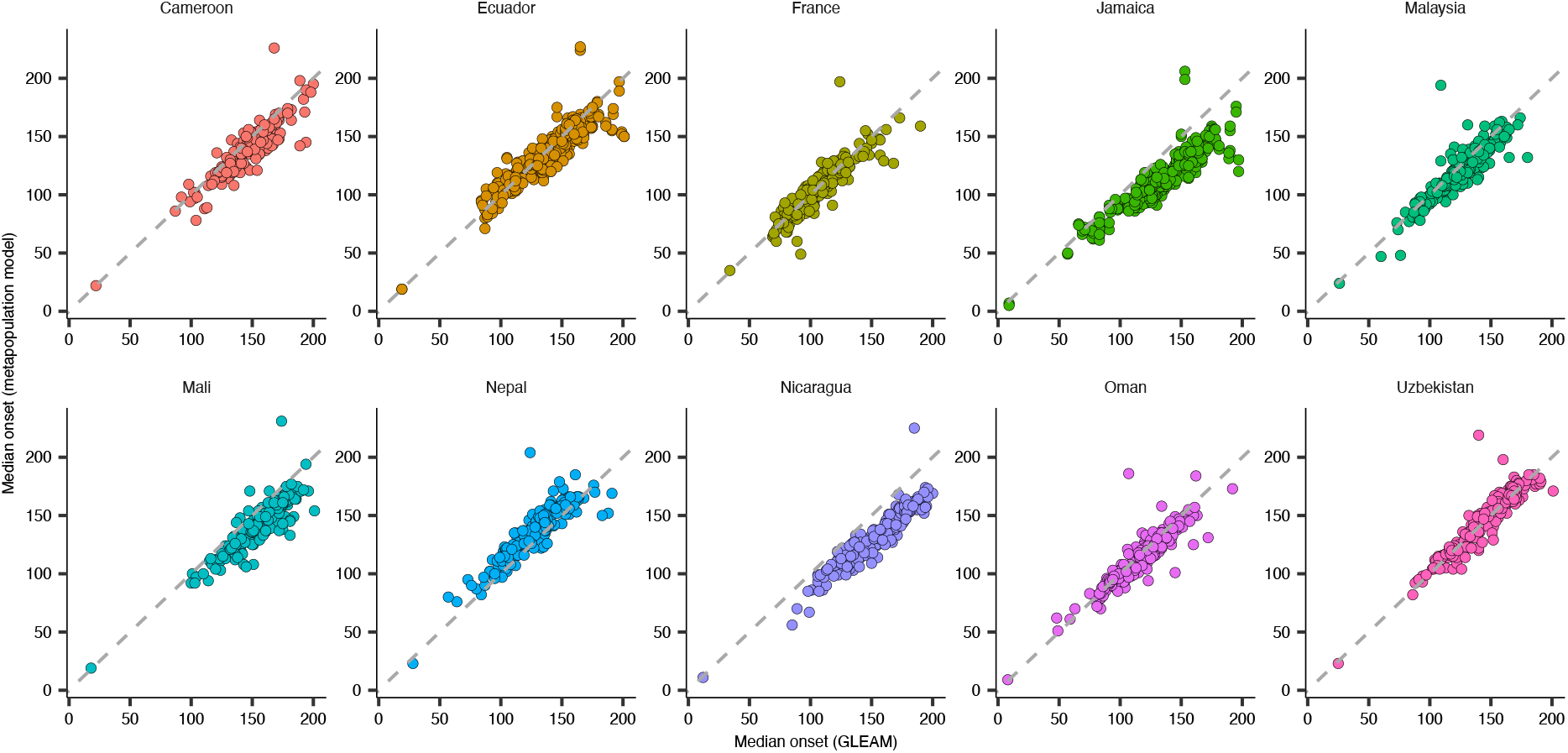
Validation of the metapopulation model against GLEAM. For ten geographically representative countries, global variant spread was simulated, initialized in the country’s capital city, in GLEAM. For each of the 10 index countries, all global countries’ epidemic onset timings as simulated using GLEAM were compared against the countries’ epidemic onset timings as simulated using the epidemic model used in this study. For both models, timings were computed as the median across 10 independent simulations. Simulations are for a variant *R*_*e*_ of 1.6.

**Extended Data Fig. 3.**
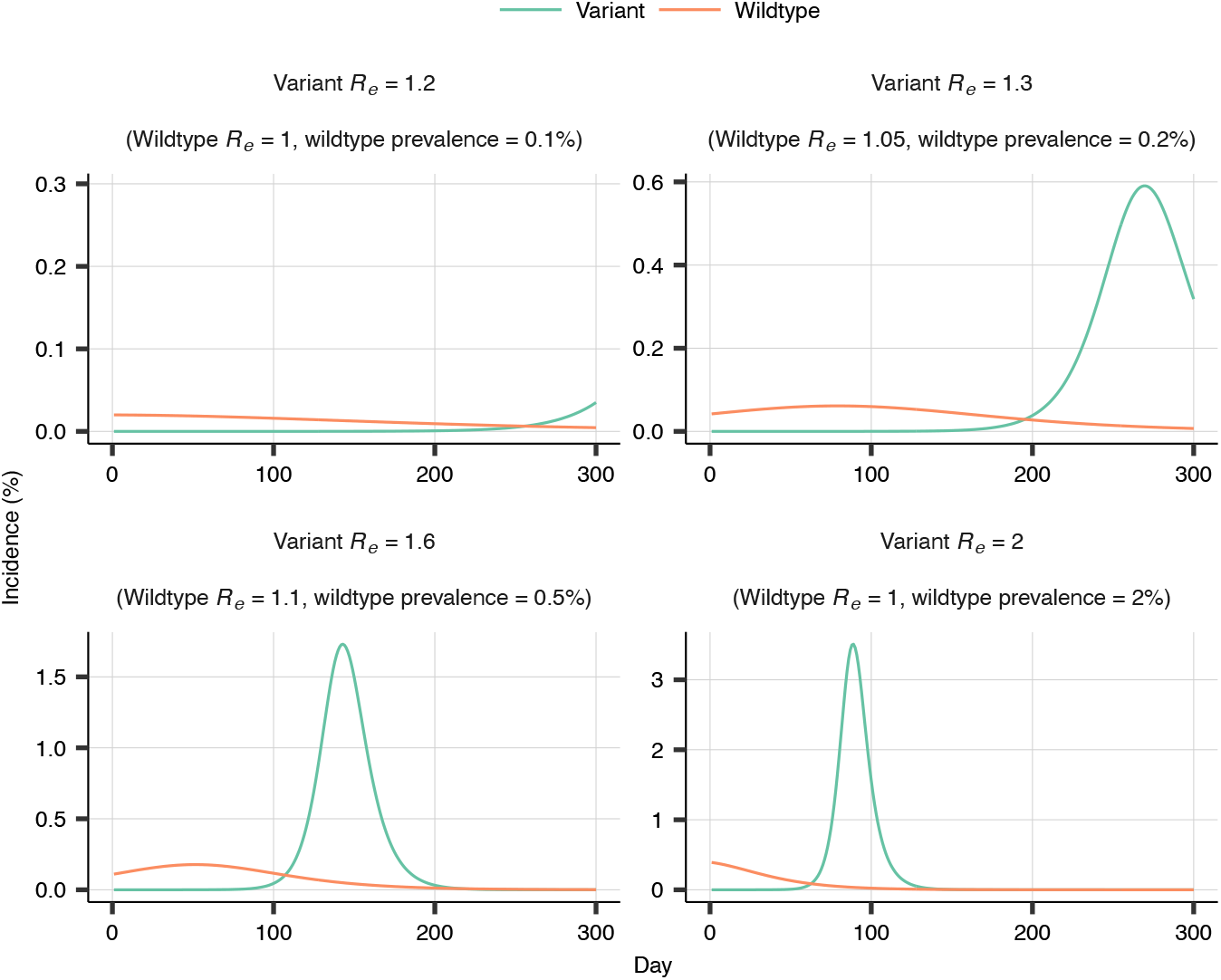
Different scenarios of variant emergence. For each of the values of variant *R*_*e*_, the corresponding panel shows the epidemiological dynamics of variant and wildtype for that scenario of variant emergence, starting from the day of variant introduction. For each value of variant *R*_*e*_, the scenario of variant emergence is characterized by a different value of wildtype *R*_*e*_ and wildtype prevalence at introduction.

**Extended Data Fig. 4.**
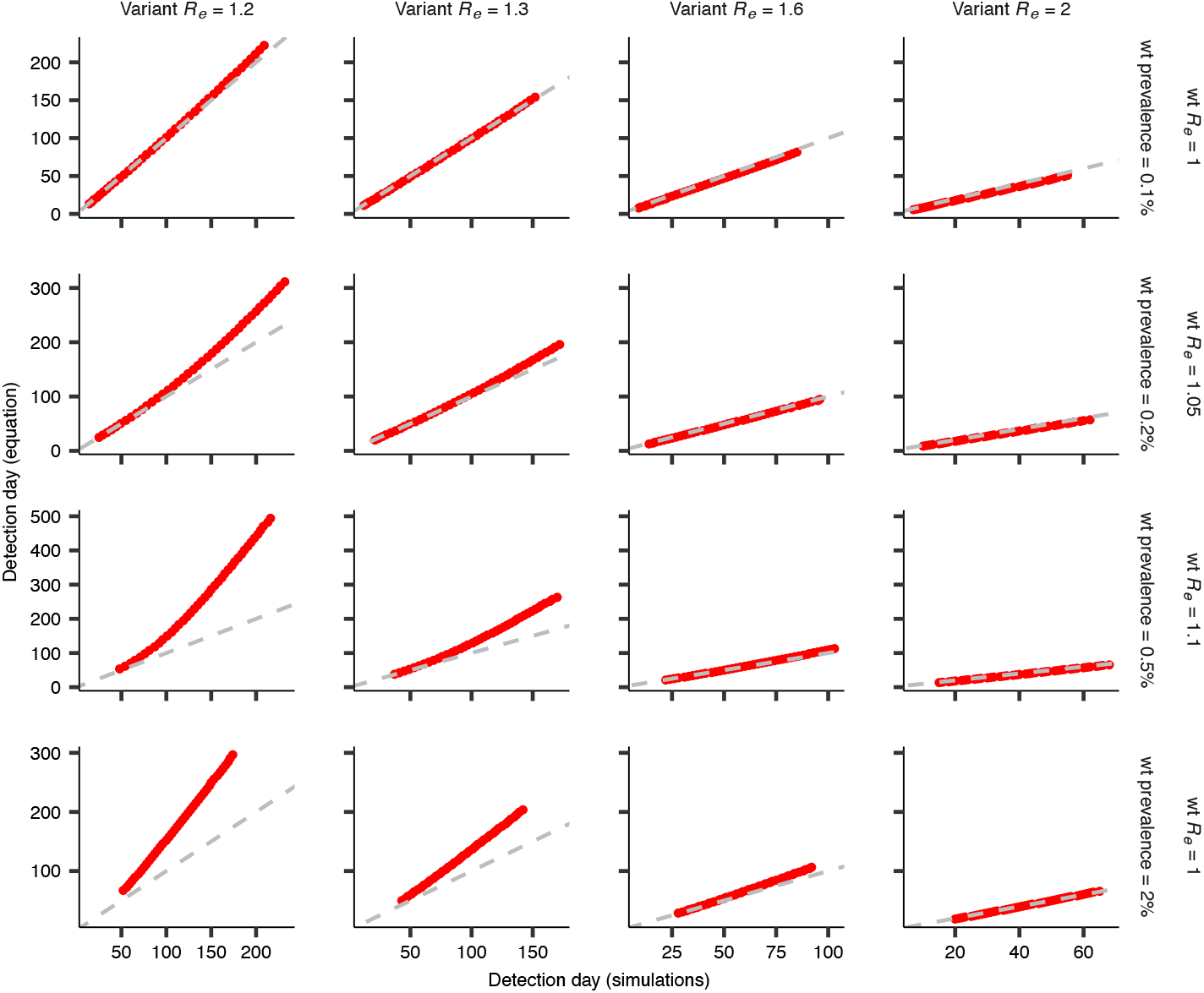
Validation of derived time to variant detection. The red points show the relationship between time to detection simulated using binomial sampling (*x-*axis) and time to detection computed using the derived mathematical model (*y*-axis). The dashed grey line corresponds to *y*=*x*. Each panel corresponds to a different variant *R*_*e*_ and different scenario of variant emergence (i.e. a different value of wildtype *R*_*e*_ and wildtype prevalence at time of variant introduction). This illustrates that the equation is valid, unless the timescales of detection are smaller than the timescales at which the logistic growth dynamics do not hold (bottom left quadrant), e.g. when susceptible depletion occurs before the variant is expected to be detected (see Methods).

**Extended Data Fig. 5.**
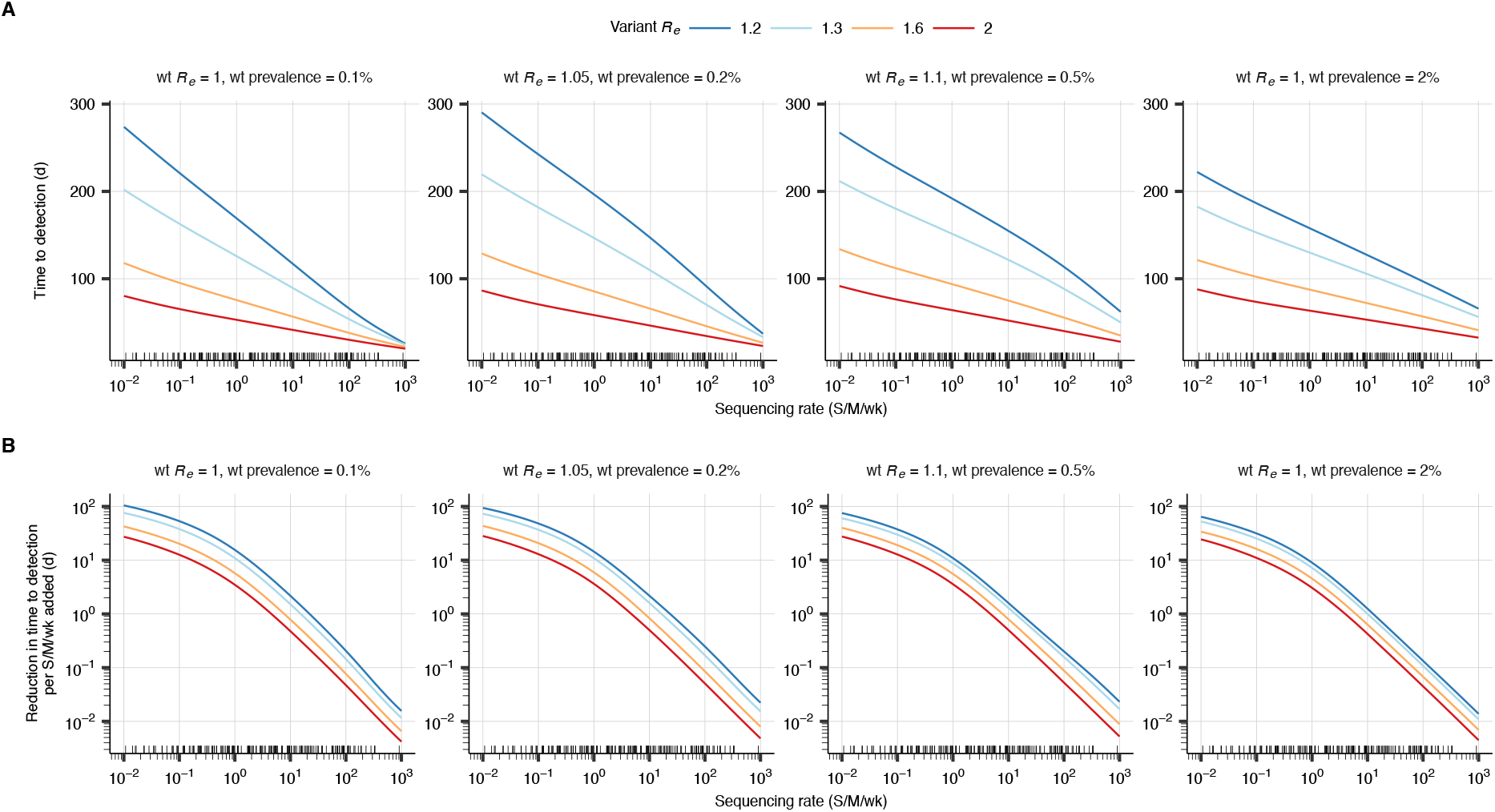
The dependence of time to variant detection on sequencing rate for varying scenario of variant emergence. **(A)** Relationship between sequencing rate and the number of days until the variant will have been detected with 95% confidence. The small black tick marks on the *x*-axes in this plot and in **B** show country-specific SARS-CoV-2 sequencing rates for 2022. Each panel corresponds to a different scenario of variant emergence, characterized by a wildtype (wt) *R*_*e*_ and wildtype prevalence at introduction. In each panel, lines are colored by value of variant *R*_*e*_. **(B)** Relationship between sequencing rate and the reduction in time to variant detection that results from increasing the existing sequencing rate (*x*-axis) by 1 S/M/wk.

**Extended Data Fig. 6.**
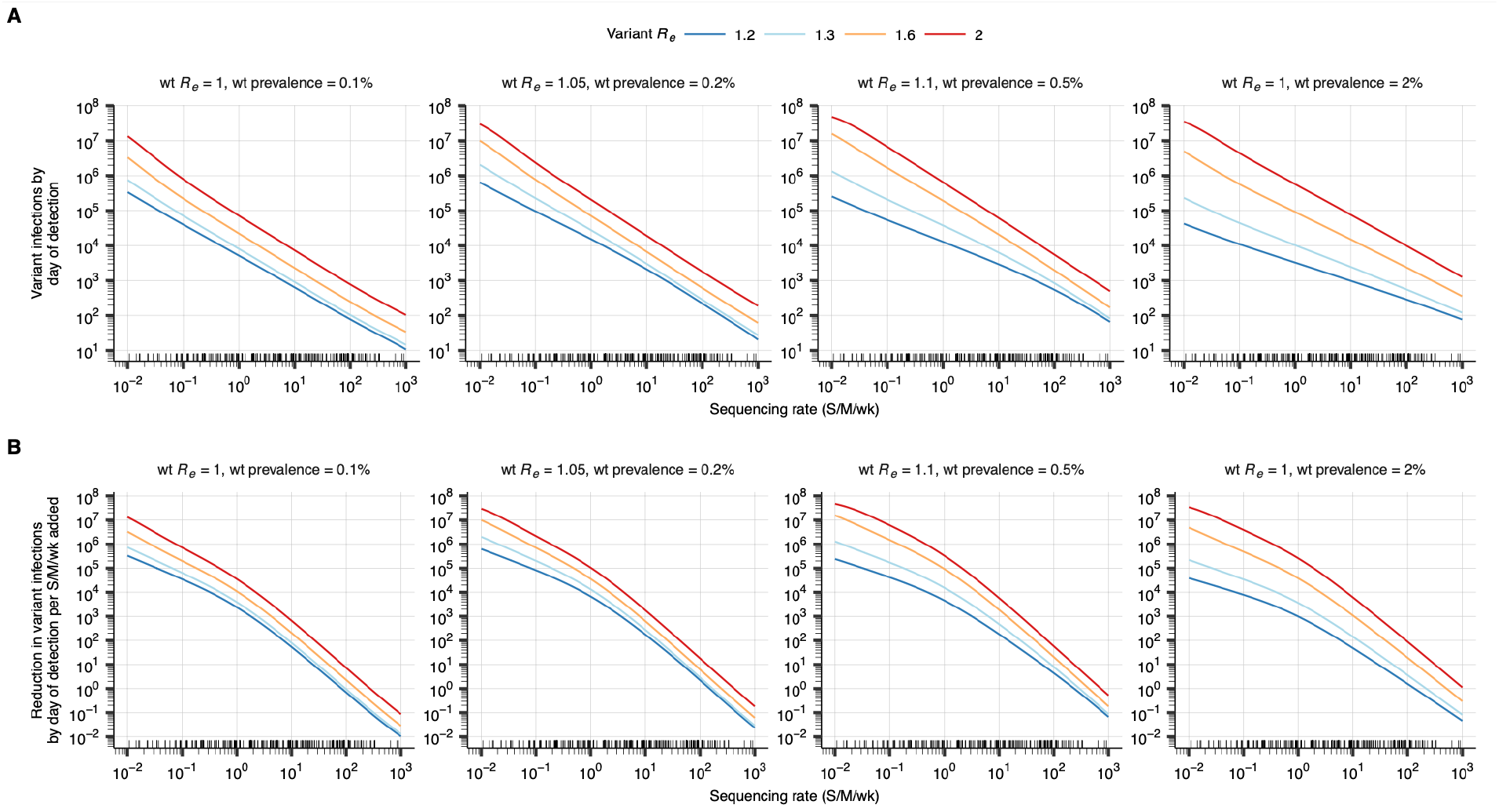
The dependence of the number of variant infections by day of detection on sequencing rate for varying scenario of variant emergence. **(A)** Relationship between sequencing rate and the number of infections by the day the variant will have been detected with 95% confidence. The small black tick marks on the *x*-axes in this plot and in **B** show country-specific SARS-CoV-2 sequencing rates for 2022. Each panel corresponds to a different scenario of variant emergence, characterized by a wildtype (wt) *R*_*e*_ and wildtype prevalence at introduction. In each panel, lines are colored by value of variant *R*_*e*_. **(B)** Relationship between sequencing rate and the reduction in the number of variant infections by the day of variant detection that results from increasing the existing sequencing rate (*x*- axis) by 1 S/M/wk.

**Extended Data Fig. 7.**
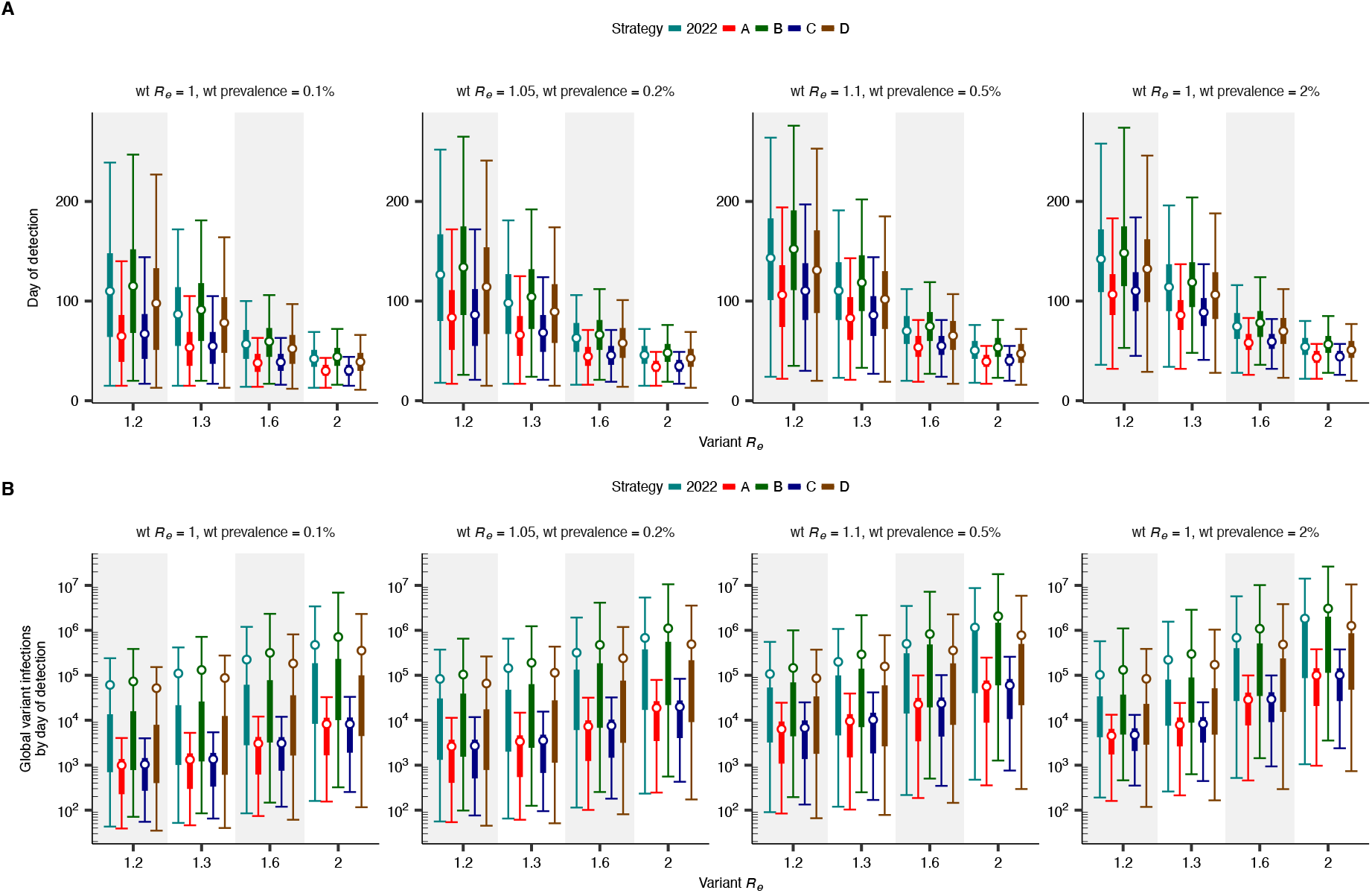
Time to global variant detection by strategy and scenario of variant emergence. **(A)** Time to global variant detection by strategy for the global distribution of respiratory virus surveillance infrastructure, by variant *R*_*e*_, for varying scenario of variant emergence (characterized by wildtype (wt) *R*_*e*_ and wildtype prevalence (*N* = 10,000 for each). Thin and thick lines correspond to 95% and 50% CIs, respectively. Points correspond to means. **(B)** Number of global variant infections by the day of first detection by strategy for the global distribution of respiratory virus surveillance infrastructure, by variant *R*_*e*_, for varying scenario of variant emergence (characterized by wildtype (wt) *R*_*e*_ and wildtype prevalence).

**Extended Data Fig. 8.**
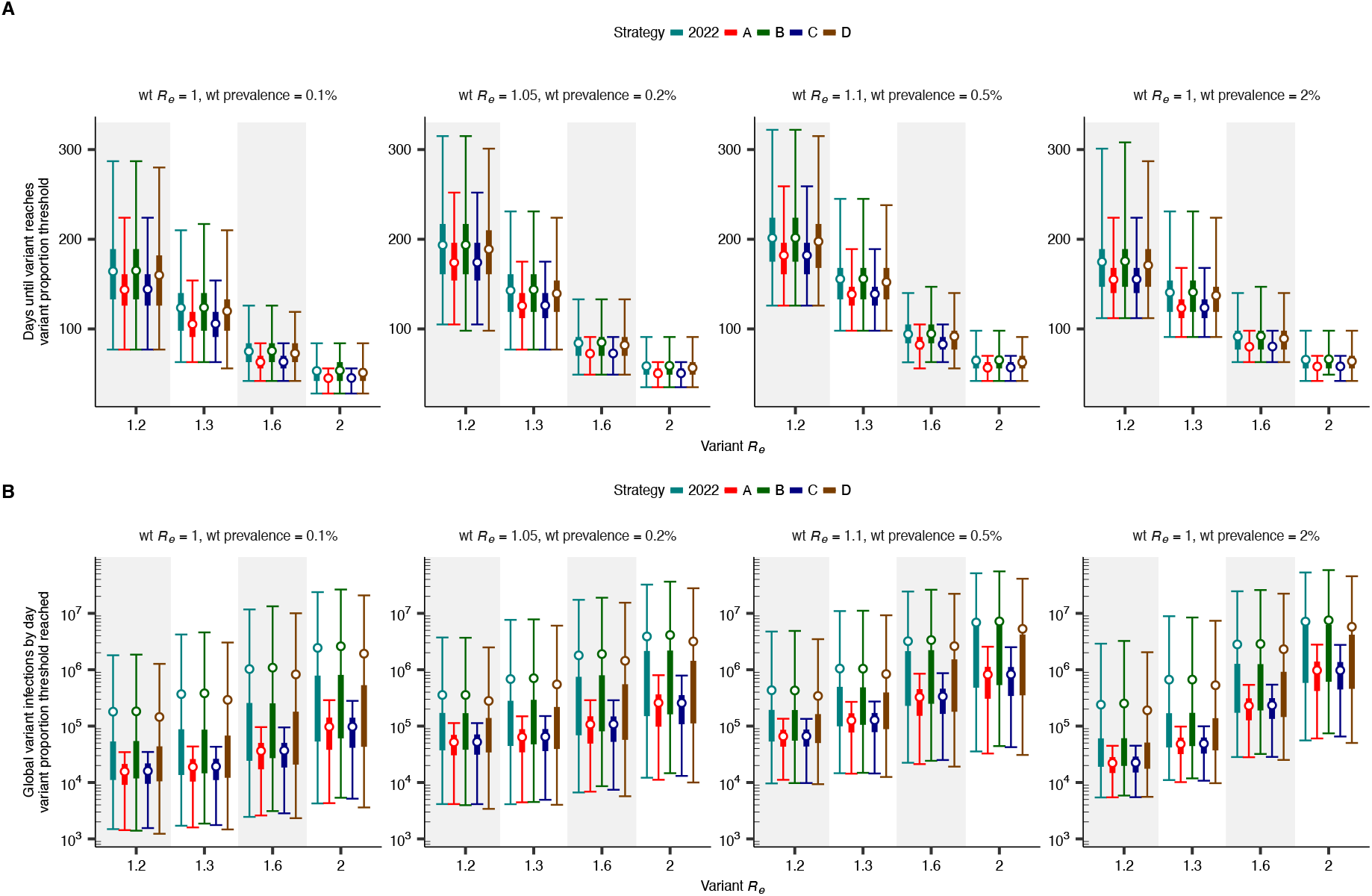
Time to ascertainment of substantial variant proportion by strategy and scenario of variant emergence. **(A)** Time until the variant was found to account for a substantial proportion of circulating virus in at least one country, by strategy for the global distribution of respiratory virus surveillance infrastructure, by variant *R*_*e*_, for varying scenario of variant emergence (characterized by wildtype (wt) *R*_*e*_ and wildtype prevalence). Thin and thick lines correspond to 95% and 50% CIs, respectively. Points correspond to means. **(B)** Number of global variant infections by the day the variant was found to account for a substantial proportion of circulating virus in at least one country, by strategy for the global distribution of respiratory virus surveillance infrastructure, by variant *R*_*e*_, for varying scenario of variant emergence (characterized by wildtype (wt) *R*_*e*_ and wildtype prevalence).

**Extended Data Fig. 9.**
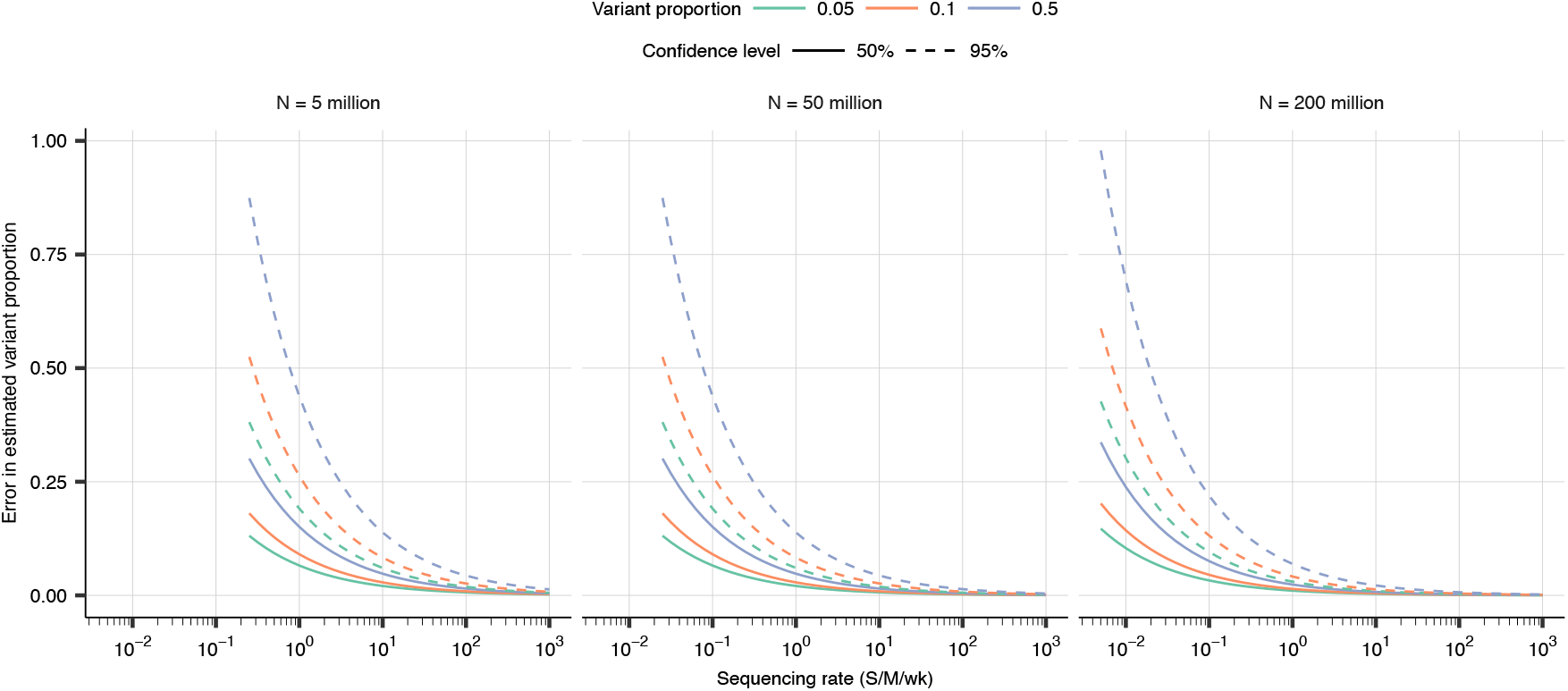
The relationship between sequencing rate and error in estimated variant proportion. For each sequencing rate given on the *x*-axis, for varying true variant proportion and population size, the *y*-axis shows the maximum error in the estimated weekly proportion of total infections attributable to the variant. This maximum error is presented for varying confidence (i.e. the *y*-axis represents the error that the error in the estimated variant proportion relative to the true variant proportion will be smaller than *n*% of the time, for *n* given by the confidence level).

**Extended Data Fig. 10.**
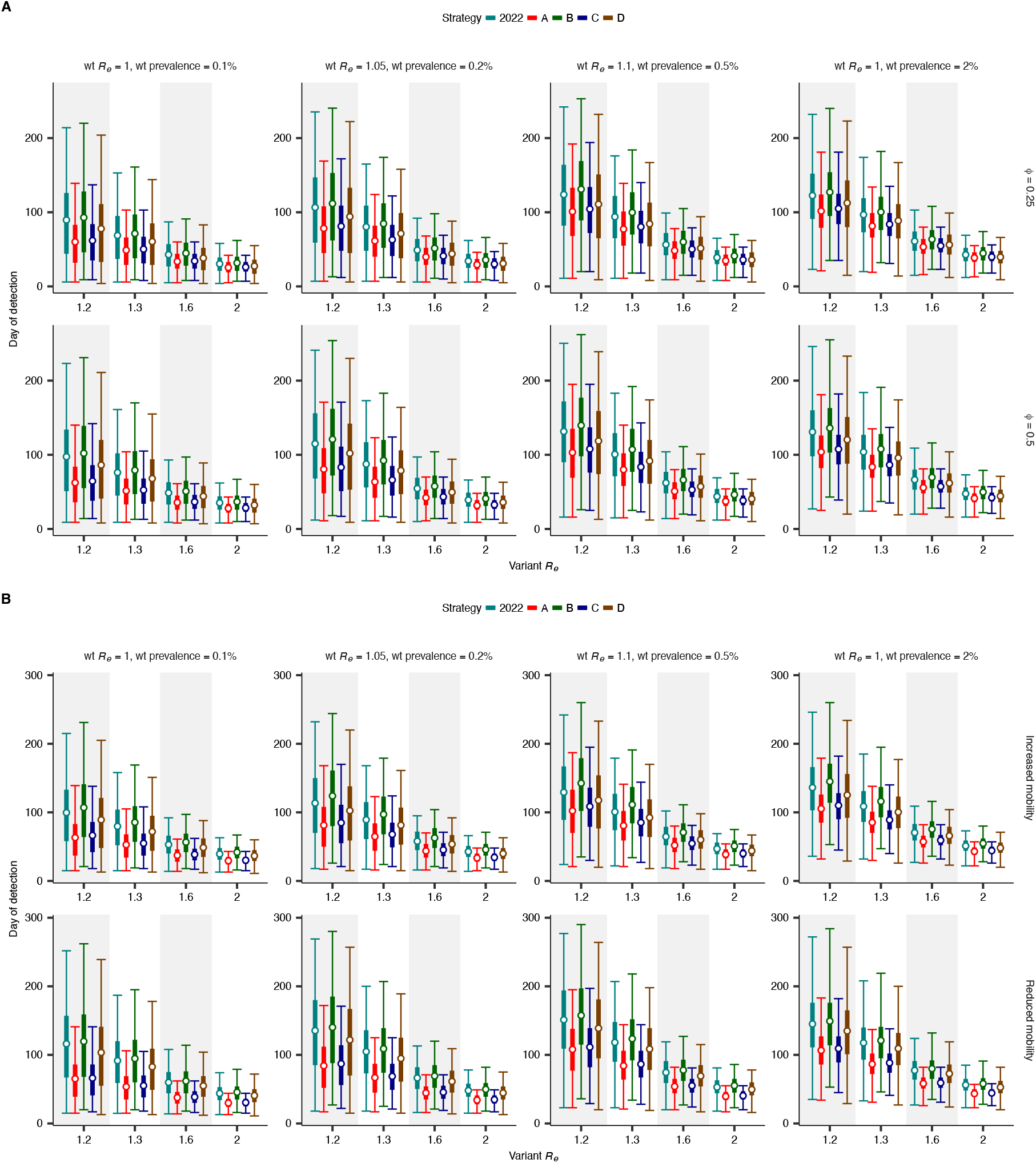
Sensitivity analyses for time to detection. **(A)** Sensitivity analysis for time to GISAID submission. Time to global variant detection by strategy for the global distribution of respiratory virus surveillance infrastructure, by variant *R*_*e*_, for varying scenario of variant emergence (characterized by wildtype (wt) *R*_*e*_ and wildtype prevalence). Thin and thick lines correspond to 95% and 50% CIs, respectively. Points correspond to means. Given a sequence in GISAID’s computed turnaround time *T*, a sequence’s adjusted turnaround time 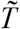 was equal to *ϕT*. These adjusted turnaround times were used to inform country-specific sequencing infrastructure in the global genomic surveillance simulations. **(B)** Sensitivity analysis for mobility rate. Time to global variant detection by strategy for the global distribution of respiratory virus surveillance infrastructure, by variant Re, for varying scenario of variant emergence (characterized by wildtype (wt) Re and wildtype prevalence). Thin and thick lines correspond to 95% and 50% CIs, respectively. Points correspond to means. Each row corresponds to a modified global mobility rate (top: baseline mobility rate multiplied by 3; bottom: baseline mobility rate divided by 3).

